# Dementia Risk Scores, *APOE,* and risk of Alzheimer disease: one size does not fit all

**DOI:** 10.1101/2024.04.27.24306486

**Authors:** Shea J. Andrews, Ana I. Boeriu, Michael E. Belloy, Alan E. Renton, Brian Fulton-Howard, Willa D. Brenowitz, Kristine Yaffe, the Alzheimer’s Disease Neuroimaging Initiative

**Affiliations:** Department of Psychiatry and Behavioral Sciences, University of California San Francisco, 505 Parnassus Ave, San Francisco, CA, USA, 94143; Department of Neurology and Neurological Sciences, Stanford University, Stanford, CA, USA, 94304; NeuroGenomics and Informatics Center, Washington University School of Medicine, St.Louis, MO, USA, 63108; Department of Neurology, Washington University School of Medicine, St.Louis, MO, USA, 63108; Department of Genetics and Genomic Sciences, Icahn School of Medicine at Mount Sinai, 1428 Madison Ave, New York, NY, USA, 10029; Kaiser Permanente Center for Health Research, 3800 N Interstate Ave, Portland, OR, USA, 97227; Department of Epidemiology and Biostatistics, University of California San Francisco, 505 Parnassus Ave, San Francisco, CA, USA, 94143; Department of Neurology, University of California San Francisco, 505 Parnassus Ave, San Francisco, CA, USA, 94143

**Keywords:** dementia, APOE, race/ethnicity, dementia risk scores

## Abstract

**Introduction:** Evaluating the generalizability of dementia risk scores, primarily developed in non-Latinx White (NLW) participants, and interactions with genetic risk factors in diverse populations is crucial for addressing health disparities.

**Methods:** We analyzed the association of the Cardiovascular Risk Factors, Aging, and Incidence of Dementia (CAIDE) and modified CAIDE (mCAIDE) scores with dementia risk using logistic regression models stratified by race/ethnicity in NACC and ADNI, and assessed their interaction with *APOE*.

**Results:** Higher CAIDE scores were associated with an increased risk of dementia in Asian, Latinx, and NLW participants but not in Black participants. In contrast, higher mCAIDE scores were also associated with an increased risk of dementia in Black participants. Unfavorable mCAIDE risk profiles exacerbated the *APOE*\*ε4 risk effect and attenuated the *APOE*\*ε2 protective effect.

**Discussion:** Our findings underscore the importance of evaluating the validity of dementia risk scores in diverse populations for their use in personalized medicine approaches to promote brain health.

## Background

Older Black and Latinx individuals are disproportionately more likely than older non-Latinx Whites (NLW) to develop Alzheimer’s disease (AD) or other related dementias [1]. Addressing health disparities in AD will require identifying individuals at risk of dementia and developing personalized disease prevention strategies [2]. As AD is a complex multifactorial neurodegenerative disease, it is essential to develop integrative risk models that combine genetic and environmental risk factors for predicting the risk of developing AD [3,4]. However, as most research examining genetic and environmental risk factors has been conducted in NLW populations, personalized medicine approaches applied to minoritized populations may not be generalizable and further exacerbate existing health disparities in AD outcomes [5].

Modifiable risk factors substantially contribute to AD risk, with up to 45% of AD cases attributable to 14 modifiable risk factors, including education, hearing loss, traumatic brain injury, hypertension, alcohol consumption, obesity, smoking, depression, social isolation, physical inactivity, air pollution, diabetes, LDL cholesterol, and visual loss [6,7]. The identification of modifiable risk factors for dementia has informed the development of dementia risk scores that are weighted composites of clinical and lifestyle risk factors that reflect the likelihood of developing dementia [4]. Dementia risk scores can be used for AD risk stratification, to facilitate communication of risk to patients, and to prioritize actionable interventions for modifiable risk factors [4].

The Cardiovascular Risk Factors, Aging, and Incidence of Dementia (CAIDE) risk score is the most widely investigated dementia risk score and has been used for enrolling participants into multi-domain intervention trials [4,8,9]. It was developed in a Finnish population-based cohort to estimate 20-year dementia risk based on an individual’s midlife risk factor profile, including age, sex, education, systolic blood pressure, body mass index, total cholesterol, and physical activity. The CAIDE risk score has good predictive accuracy for AD within the population in which it was developed (AUC = 0.75 - 0.78), with individuals in the highest risk profile having a 29-35% increased risk of developing dementia [12]. However, the prognostic utility of CAIDE in other populations has been more limited [9–11].

The lack of generalizability of CAIDE in other populations may reflect underlying differences in the risk factors associated with dementia pathogenesis, or, where the specific combination of predictors is appropriate across populations, the weights assigned to each risk factor may need to be recalibrated in different populations [10]. As such, CAIDE has recently been recalibrated to develop a modified CAIDE (mCAIDE) risk score based on a multi-ethnic cohort of community-dwelling older adults in the US to predict late-life dementia that reweights age and education to account for the older age group and higher educational attainment compared to the original development population [12]. The mCAIDE demonstrated good discriminative performance between controls and all-cause dementia (AUC = 0.8); however, further external validation and comparison to CAIDE is required [12].

In addition to modifiable risk factors, the *APOE*\*ε4 allele is the strongest genetic risk factor for late-onset AD, while the *ε2 allele is associated with a reduced risk of AD [13]. Two versions of the CAIDE risk score were initially derived, one excluding *APOE* and one including *APOE*, while the mCAIDE did not include *APOE* due to including only readily assessable and self-reported measurements. However, *APOE* exhibits ancestry-specific effects, with the *ε4 risk effect attenuated in participants of African and Amerindian ancestry [14,15]. This attenuation may be due to gene-environment interactions whereby genetic differences in disease risk are more influential in positive social environments, allowing the underlying genetic predisposition to emerge more distinctly [16,17]. To date, research investigating the moderating effect of dementia risk scores on the association between *APOE* and dementia or cognitive impairment has produced mixed results, with few studies evaluating the effect across racial/ethnic groups [18–23].

Due to the under-representation of minoritized populations in AD genetic and epidemiological studies, it is critical to determine the generalizability of dementia risk scores across populations and determine to what extent they moderate genetic liability for dementia. To address this knowledge gap, we evaluated the association of the CAIDE and mCAIDE risk scores with all-cause dementia and to what extent they moderate the association of *APOE* with dementia across NLW, Black, Latinx, and Asian Americans.

## Methods

### Participants

This cross-sectional case-control study uses data from two cohorts – the National Alzheimer’s Coordinating Center Uniform Dataset (NACC UDS) and the Alzheimer’s Disease Neuroimaging Initiative (ADNI). The NACC UDS consists of over 45,000 participants from 30+ past and present US-based Alzheimer’s Disease Core Centers and Alzheimer Disease Research Centers funded by the National Institute on Aging [24]. ADNI was launched in 2003 as a public-private partnership with the primary goal of testing whether serial magnetic resonance imaging (MRI), positron emission tomography (PET), other biological markers, and clinical and neuropsychological assessment can be combined to measure the progression of mild cognitive impairment (MCI) and early AD [25].

Race and ethnicity were self-reported by study participants, with categories defined by the National Institutes of Health, including American Indian or Alaska Native, Asian, Black or African American, Native Hawaiian or Other Pacific Islander, and White. Ethnicity categories included Hispanic or Latino or not Hispanic or Latino. If individuals did not identify with these racial and ethnic categories, they could report “other.” We analyzed baseline visit data for non-Latinx White, Black, Latinx, and Asian participants who were at least age 55 at their initial visit or whose estimated age-of-onset of cognitive impairment was at least 55, had *APOE* genotyping data, were cognitively unimpaired or had a primary diagnosis of MCI or all-cause dementia. Diagnostic criteria for NACC and ADNI have been previously described [24,25]. Participants with autosomal dominant AD or FTD mutations were excluded.

### CAIDE Risk Score

The CAIDE and mCAIDE risk scores for each participant were calculated using the published equations using the following variables: age, sex, hypertension, obesity, and hypercholesteremia (Supplementary Tables 1 & 2) [8,12]. Physical activity assessments were unavailable; however, CAIDE remains predictive of dementia when physical activity is not included [26]. The CAIDE risk score includes *APOE* genotype in its algorithm, however, as the mCAIDE does not include *APOE* and due to the observed ancestry-specific effects of *APOE* on AD, we did not include *APOE* in the estimation of CAIDE [15]. Supplementary Tables 1 & 2 show the scoring algorithm for each risk factor, with the sum of points across risk factors representing the total CAIDE/mCAIDE score. The CAIDE score uses age and education cutoffs of <47, 47-53, >53 and ≥10, 7-9, <7 years, respectively. In contrast, the mCAIDE applies age cutoffs of <65, 56-72, >73 years, and education levels of ≥16, 12-16, and <12 years. In NACC and ADNI, we utilized self-reported data for age, sex, and educational attainment. Obesity was defined as a Body mass index (BMI) >30, and hypertension as a sitting systolic blood pressure >140mmHg. For NACC, hypercholesteremia was identified through self-reported medical history or clinician assessment. In ADNI, it was determined by a fasting total cholesterol level exceeding 6.21 mmol/L. Missing values were observed in BMI (8.68%), hypercholesterolemia (6.59%), education (0.62%), and hypertension (0.28%). To address this, missing data was imputed using a Random Forrest algorithm via the ‘MissForest’ R package, package, which implements a non-parametric method for imputing missing values for both continuous and categorical data simultaneously within a multiple imputation framework [27]. The CAIDE and mCAIDE scores were standardized to have a mean of 0 and standard deviation of 1 and categorized into tertiles representing favorable (CAIDE < 5; mCAIDE < 2), intermediate (CAIDE => 5 & < 9; mCAIDE => 3 & < 7), and unfavorable (CAIDE => 9; mCAIDE => 7) risk profiles.

### *APOE* Genotyping

*APOE* haplotypes for NACC were determined from the single-nucleotide variants rs7412 and rs42935848 and for ADNI from pyrosequencing of *APOE* codons 112 and 158 [28,29]. *APOE* haplotypes were combined into three groups: ε2+ (ε2/ε2, ε2/ε3), ε4+ (ε2/ε4, ε3/ε4, ε4/ε4) and ε3/ε3.

### Statistical analysis

Baseline characteristics of the joint NACC and ADNI cohorts were summarized across racial/ethnic groups as percentages for categorical variables and mean and SD for continuous variables and racial/ethnic differences determined using ANOVA and Chi-square tests (Table 1). Descriptive statistics by cognitive status and race/ethnicity are presented in Supplementary Table 3. Multivariate logistic regression models stratified by race/ethnicity were used to evaluate the association between *APOE* genotype and standardized CAIDE/mCAIDE risk scores with ADRD/MCI. To compare effect sizes across racial/ethnic groups, we used the z-score method, where the differences between group-specific beta coefficients were standardized by their combined standard errors, yielding z-scores (z = [b - b]/sqrt[SE ^2^ + SE ^2^]) [30]. These z-scores were then used to calculate two-tailed p-values to assess the statistical significance of the differences observed. The Area Under the Curve (AUC) was used to further evaluate the discriminative ability of the CAIDE and mCAIDE risk scores across race/ethnicity, with Delong’s statistical test used to compare performance across different models. To determine if modifiable risk profiles moderated the association of *APOE* genotype with ADRD/MCI, we evaluated interactions on the additive and multiplicative scales. On the additive scale, we combined *APOE* and CAIDE/mCAIDE risk categories (9 categories with intermediate risk profiles, *APOE* ε3/ε3 as the reference category). We then used logistic regression models to evaluate the association of the combined *APOE* CAIDE/mCAIDE risk categories with ADRD/MCI. On the multiplicative scale, we introduced an interaction term between CAIDE/mCAIDE and *APOE* genotype within the logistic regression models.

**Table 1:** Cohort description.

|  | NLW<br>N = 16,962 | Asian<br>N = 573 | Black<br>N = 2,259 | Latinx<br>N = 961 | P |
| --- | --- | --- | --- | --- | --- |
| **Cohort** |  |  |  |  | - |
| NACC | 14,992 (88%) | 520 (91%) | 2,107 (93%) | 868 (90%) |  |
| ADNI | 1,970 (12%) | 53 (9.2%) | 152 (6.7%) | 93 (9.7%) |  |
| **Female** | 9,076 (54%) | 324 (57%) | 1,647 (73%) | 614 (64%) | 4.7e-71 |
| **Age** | 73 (8) | 72 (8) | 73 (8) | 72 (8) | 1e-05 |
| **Education** | 15.96 (2.82) | 16.01 (3.56) | 14.36 (3.26) | 12.80 (4.96) | <1e-100 |
| **BMI** | 26.6 (4.7) | 24.1 (3.5) | 29.3 (5.9) | 27.6 (4.8) | <1e-100 |
| **Hypertension** | 7,510 (44%) | 287 (50%) | 1,616 (72%) | 528 (55%) |  |
| **Hypercholesterolemia** | 7,652 (45%) | 281 (49%) | 1,114 (49%) | 499 (52%) |  |
| **CAIDE** | 6.49 (1.79) | 6.39 (1.71) | 7.10 (1.90) | 7.36 (2.31) | 9e-85 |
| High (10-14) | 2,644 (16%) | 69 (12%) | 462 (20%) | 282 (29%) |  |
| Mid (5-9) | 11,392 (67%) | 399 (70%) | 1,526 (68%) | 553 (58%) |  |
| Low (0-4) | 2,926 (17%) | 105 (18%) | 271 (12%) | 126 (13%) |  |
| **mCAIDE** | 4.44 (2.06) | 4.18 (2.01) | 5.12 (2.13) | 5.10 (2.28) | 2.4e-64 |
| High (7-11) | 2,875 (17%) | 77 (13%) | 609 (27%) | 272 (28%) |  |
| Mid (3-6) | 10,987 (65%) | 369 (64%) | 1,393 (62%) | 566 (59%) |  |
| Low (0-2) | 3,100 (18%) | 127 (22%) | 257 (11%) | 123 (13%) |  |
| **APOE** |  |  |  |  | 4.8e-32 |
| ε2+ | 1,526 (9.0%) | 66 (12%) | 284 (13%) | 49 (5.1%) |  |
| ε3/ε3 | 8,385 (49%) | 345 (60%) | 916 (41%) | 562 (58%) |  |
| ε4+ | 7,051 (42%) | 162 (28%) | 1,059 (47%) | 350 (36%) |  |
| **Diagnosis** |  |  |  |  | 1.3e-27 |
| CU | 8,189 (48%) | 293 (51%) | 1,377 (61%) | 446 (46%) |  |
| MCI | 3,296 (19%) | 125 (22%) | 333 (15%) | 206 (21%) |  |
| ADRD | 5,477 (32%) | 155 (27%) | 549 (24%) | 309 (32%) |  |

We conducted several sensitivity analyses to evaluate the robustness of our models. First, we also examined the association of *APOE*, CAIDE/mCAIDE, and their combination with ADRD only, AD/MCI, and AD only. Second, we evaluated the association of individual risk factors with ADRD/MCI. Third, we conducted sex-stratified sensitivity analyses to evaluate the intersectional effect of sex and race on the association of CAIDE/mCADIE risk scores (excluding sex) with risk of ADRD/MCI. Finally, we evaluated the interaction between *APOE* and an mCAIDE score composed only of modifiable risk factors (m^2^CAIDE; education, hypertension, obesity, and hypercholesteremia) with ADRD/MCI to determine if the observed interactions are independent from age and sex.

Results are reported as odds ratios and 95% confidence intervals (OR [95% CI]). P-values were 2-sided with statistical significance set at less than 0.05. All analyses were performed using R version 4.2.2.

## Results

### Participant Characteristics

A total of 20,755 older adults were included in this analysis (aged 73 ± 8 years; 56% Female; 82% NLW, 11% Black, 4.6% Latinx, and 2.8% Asian). Heterogeneity with respect to age, education, gender, hypertension, hypercholesteremia, BMI, clinical diagnosis, and *APOE* genotype were present between racial/ethnic groups (Table 1; Supplementary Tables 4 & 5).

### Higher CAIDE scores are associated with increased risk of MCI/ADRD in NLW, Latinx, and Asian participants, but not among Black participants

Among all participants, a one standard deviation increase in CAIDE was significantly associated with 15% higher odds of ADRD/MCI (OR [95%CI] = 1.15 [1.12, 1.18], p = 1.1e-21). In race/ethnicity-stratified analyses, CAIDE was associated with 45%, 22%, and 16% higher odds of ADRD/MCI in Asian, Latinx, and NLW participants, respectively, with no significant association observed in Black participants (Figure 2, Supplementary Tables 6 & 7). The AUCs were 0.64 for NLW, 0.61 for Latinx, 0.63 for Black, and 0.67 for Asian participants (Supplementary Table 8). The magnitude of association in Asian participants was significantly higher than that of NLW, Latinx, and Black participants. Similarly, the magnitude of association was higher in NLW and Latinx participants than in Black participants. Similar findings were observed in sensitivity analyses examining the association of CAIDE, with ADRD only, AD/MCI, and AD only (Supplementary Tables 6 & 7). In sex-stratified analyses, higher CAIDE scores were associated with increased odds of MCI/ADRD in female NLW, Asian, and Latinx participants, but were non-significant in female Black participants or males in any racial/ethnic group (Figure 2; Supplementary Tables 9 & 10). Individual risk factors associated with a reduced risk of ADRD/MCI included higher education attainment and higher BMI (NLW, Black, and Latinx), while older age, male, and higher systolic blood pressure (NLW, Latinx, Black) were associated with increased risk. Hypercholesterolemia was not significantly associated with ADRD/MCI (Supplementary Table 11). AUC values were 0.7 for NLW, 0.72 for Latinx, 0.74 for Black, and 0.77 for Asian participants.

### Higher mCAIDE scores are associated with increased risk of MCI/ADRD in all populations

To assess whether using a dementia risk score developed in a US population is associated with increased odds of ADRD/MCI, we evaluated the association of mCAIDE with dementia. Among all participants, a one standard deviation increase in mCAIDE was significantly associated with 29% higher odds of dementia (OR [95%CI] = 1.29 [1.26, 1.33], p = 2.1e-67). In race/ethnicity stratified analysis, a one-standard deviation increase in mCAIDE was significantly associated with increased odds of ADRD/MCI in all populations with a step-wise reduction in the magnitude of association in Asian, Latinx, NLW, and Black participants (Figure 2, Supplementary Tables 12 & 13). Similar to CAIDE, the association was significantly stronger in Asian participants compared to NLW, Latinx, and Black participants; and also stronger in Latinx and NLW participants when compared to Black participants. These patterns remained consistent in sensitivity analyses examining the association of the CAIDE risk score with ADRD only, AD/MCI, and AD only (Supplementary Tables 12 & 13). The AUCs were 0.65 for NLW, 0.63 for Latinx, 0.64 for Black, and 0.7 for Asian participants, with mCAIDE significantly improving discriminative ability compared to CAIDE for all race/ethnic groups except for Black participants (Supplementary Table 8). In sex-stratified analyses, mCAIDE was associated with increased odds of MCI/ADRD across NLW, Asian, Black, and Latinx females, while in Males, higher mCAIDE scores were significantly associated with increased risk in NLW and Asian participants and trended towards significance in Latinx participants (Figure 2; Supplementary Tables 14 & 15).

### Unfavorable modifiable risk profiles exacerbate the risk of *APOE*\*ε4 and attenuate the protective effect of *APOE*\*ε2

In race/ethnicity stratified analyses, the *APOE\**ε4 status was associated with greater odds of ADRD/MCI in each population, while the *APOE\**ε2 status was significantly associated with reduced risk in NLW and Blacks only (Figure 1; Supplementary Table 6 & 10). When *APOE* alleles and CAIDE risk profiles were combined, unfavorable risk profiles exacerbated the risk effect of *APOE*\*ε4 and attenuated the protective effect of *APOE*\*ε2, predominantly in NLW participants (Figure 3; Supplementary Table 6). In NLW *APOE\**ε4 carriers, a favorable CAIDE profile was associated with 71% higher odds of ADRD/MCI (OR [95%CI] = 1.71 [1.5, 1.95], p = 1.6e-15), while an unfavorable risk profile was associated with nearly three times higher odds of dementia (OR [95%CI] = 2.97 [2.58, 3.41], p = 5.7e-53). Conversely, in NLW *APOE*\*ε2 carriers, a favorable CAIDE profile was associated with nearly two times lower odds of ADRD/MCI (OR [95%CI] = 0.49 [0.38, 0.63], p = 3.2e-08), while an unfavorable risk profile mitigated the protective effect of *APOE*\*ε2 (OR [95%CI] = 1.26 [0.94, 1.7], p = 0.12). In Black participants, CAIDE risk profiles did not moderate the association of *APOE* genotype with ADRD/MCI, while in Latinx and Asian participants, there was a less distinct pattern of effect moderation. On the multiplicative scale, the only significant interaction was between *APOE\**ε2 and CAIDE in Black participants (p = 0.03; Supplementary Table 16).

**Figure 1:**
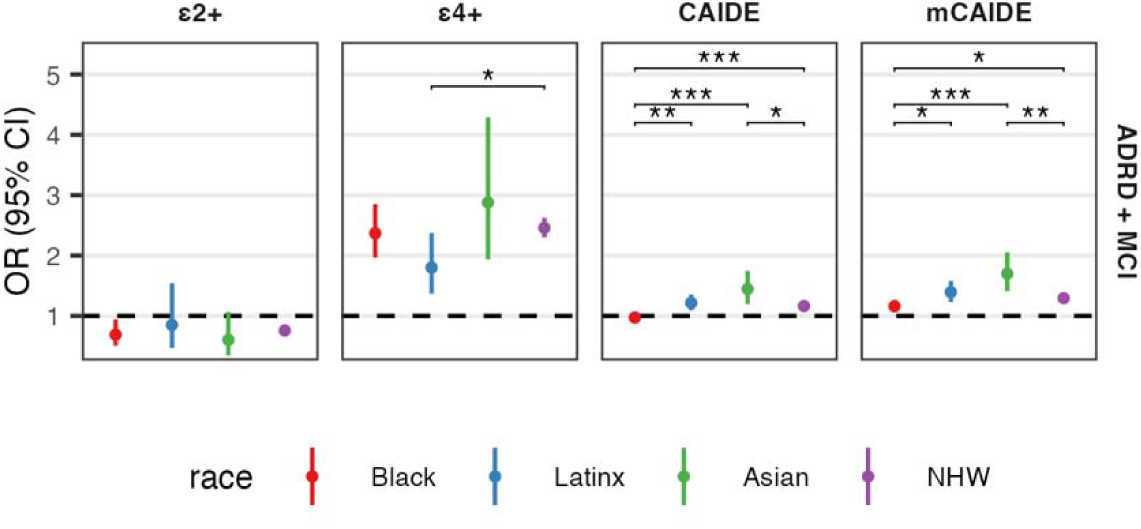
Association of *APOE* genotype, CAIDE, and mCAIDE with ADRD/MCI across race/ethnicity.

**Figure 2:**
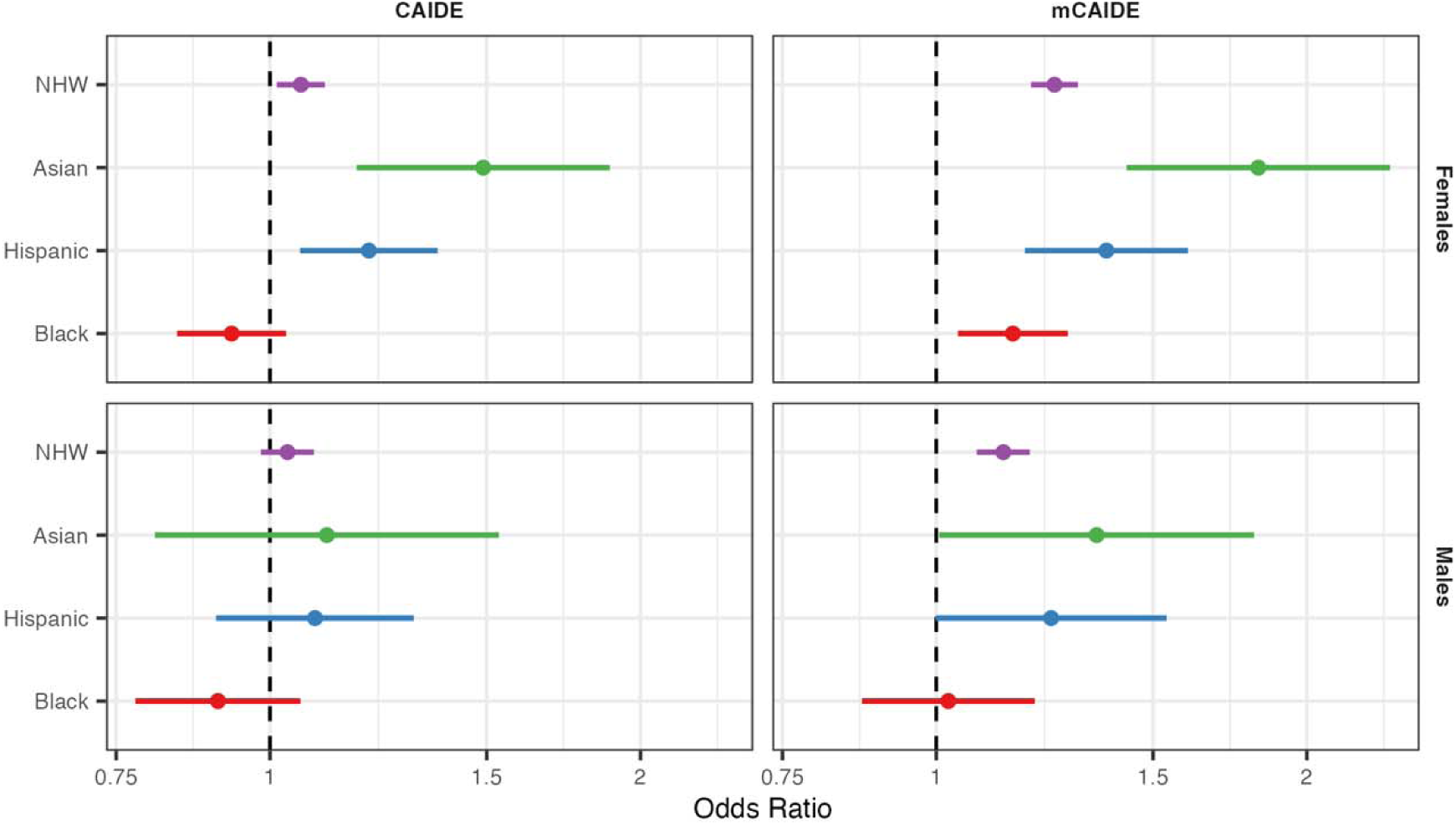
Association of CAIDE and mCAIDE risk scores with MCI/ADRD stratified by gender and race/ethnicity.

**Figure 3:**
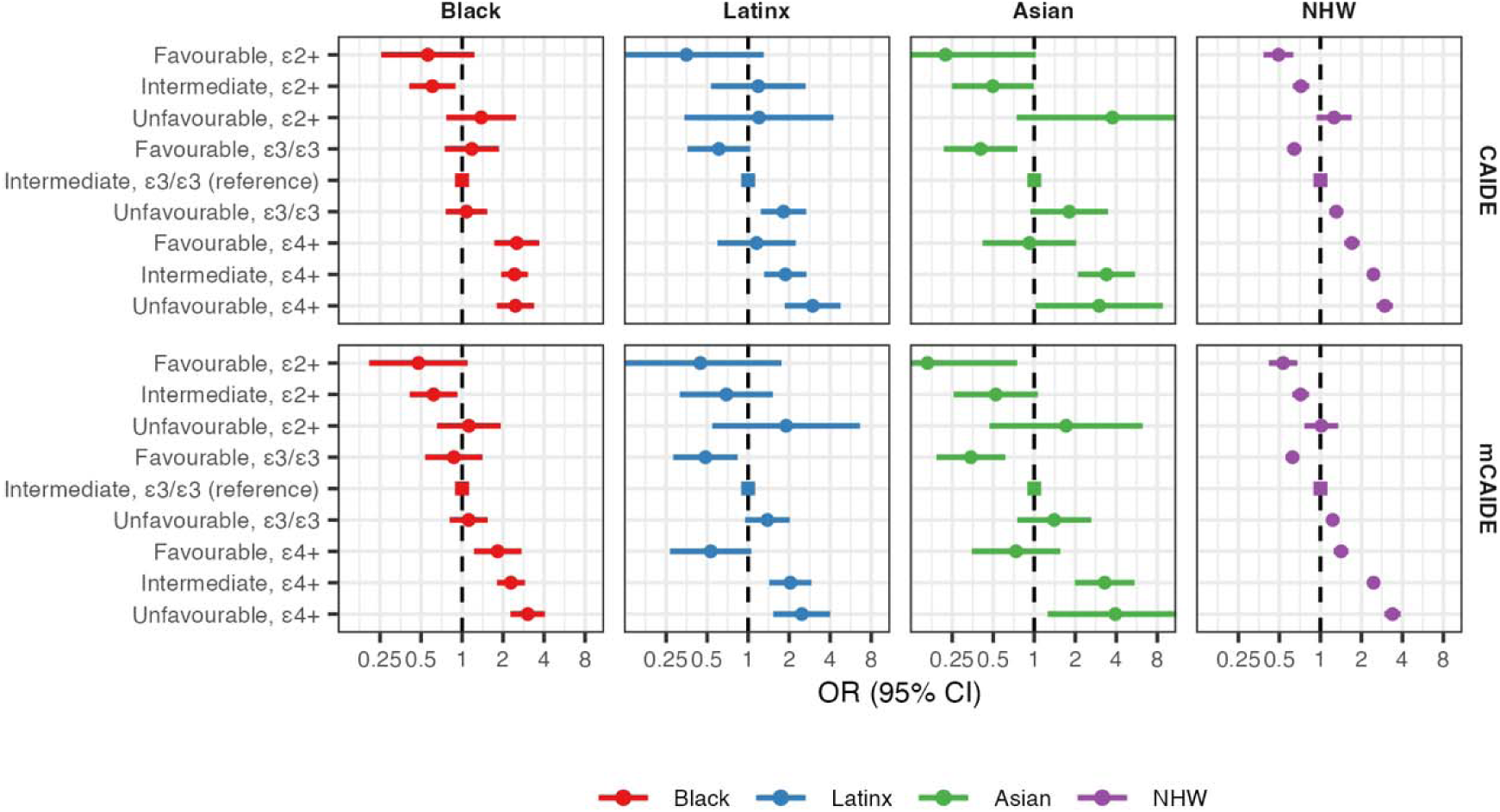
Risk of ADRD/MCI according to genetic and modifiable risk factor burden. The CAIDE and mCAIDE were categorized into tertiles representing favorable (CAIDE < 5; mCAIDE < 2), intermediate (CAIDE => 5 & < 9; mCAIDE => 3 & < 7), and unfavorable (CAIDE => 9; mCAIDE => 7) risk profiles. Intermediate risk profiles and *APOE* ε3/ε3 were used as the reference category.

When combining *APOE* alleles with mCAIDE risk profiles, a similar pattern of unfavorable risk profiles exacerbating *APOE\**ε4 risk and attenuating *APOE*\*ε2 protection was observed in NLW participants, with a less distinct pattern of effect moderation in Latinx and Asian participants (Figure 3; Supplementary Table 12). However, in contrast to CAIDE, increasingly unfavorable mCAIDE profiles exacerbated the risk effect of *APOE\**ε4 in Black participants. In sensitivity analyses, unfavorable risk profiles were similarly observed to moderate the association of *APOE* with ADRD only, AD/MCI, and AD only, though the magnitude of the effect was attenuated in AD. On the multiplicative scale, the only significant interaction between *APOE*\*ε4 and mCAIDE was in NLW participants (p=0.025; Supplementary Table 17). To determine if the observed moderation of *APOE* by mCAIDE was driven by age and/or sex, we further evaluated the effect moderation by a mCAIDE risk score composed only of modifiable risk factors (education, hypertension, obesity, and hypercholesteremia) on ADRD/MCI. Similar to our primary analyses, unfavorable modifiable only risk profiles attenuated the protective effect of *APOE*\*ε3 and exacerbated the risk effect of *APOE\**ε4 on ADRD/MCI (Supplementary Figure 1; Supplementary Table 18).

## Discussion

In this study, we found that a higher dementia risk burden assessed using the CAIDE risk score was associated with higher odds of ADRD/MCI; however, there was significant heterogeneity in the magnitude of association across racial/ethnic groups. CAIDE was associated with higher odds of ADRD/MCI in Asians, followed by Latinx and NLW, with no significant association in Blacks. However, using a modified CAIDE risk score developed to predict the risk of AD in community-dwelling older adults in the US, a higher dementia risk burden was also associated with increased risk in Blacks, though the magnitude of association was smaller than that of NLW, Asians, and Latinx. Finally, unfavorable risk profiles were observed to moderate the association of *APOE* with ADRD/MCI, such that the risk effect of *APOE\**ε4 was exacerbated, while the protective effect of *APOE*\*ε2 was attenuated. However, this pattern of association was only observed in NLWs and Blacks when using the mCAIDE.

Our results extend a limited but growing body of literature evaluating the generalizability of dementia risk scores across diverse populations. When used to predict 3-5 year incident dementia in 11,143 dementia-free individuals aged over 65 from China, Cuba, the Dominican Republic, Mexico, Peru, Puerto Rico, and Venezuela, CAIDE (excluding *APOE*) exhibited poor discriminative ability (c-statistic = 0.52 – 0.63) [10]. In a population-based multi-ethnic US cohort (41% NLW, 11% Chinese American, 26% African American, 21% Latinx) of 4,392 middle-aged and older adults, baseline CAIDE risk scores (including *APOE*) were associated with worse global cognition, processing speed, and working memory 10 years later [11]. Compared to NLW, the magnitude of association between CAIDE and global cognition was greater in Latinx and African Americans, but not in Chinese Americans. In a rural community-dwelling cohort of NLW and Latinx middle and older-aged adults, CAIDE (including and excluding *APOE*) was associated with worse global cognition and the strength of association differing by racial/ethnic group [31]. These results, and those reported here, highlight that the CAIDE risk score exhibits racial/ethnic-specific associations.

The racial/ethnic differences in the association of CAIDE with dementia and cognitive performance, likely reflect differences in sample and methodological characteristics between the original development study and subsequent cohorts. In particular, CAIDE was developed in a highly homogenous sample to predict the mid-life risk of dementia, making it less generalizable to more diverse samples. The lack of generalizability across populations may reflect underlying differences in the risk factors associated with dementia pathogenesis [32]. This highlights the need to optimize the best combination of predictors for constructing dementia risk scores. Alternatively, where the specific combination of predictors is appropriate across populations, the weighting assigned to each risk factor may need to be recalibrated when applied to different populations. As such, the mCAIDE risk score was developed to predict late-life dementia by recalibrating the CAIDE risk score to better reflect US demographics, including age and educational attainment [12].

We used the mCAIDE to determine if reweighting of risk factors used in the calculation of CAIDE would modify the association with dementia [12]. While we still observed racial/ethnic differences in the magnitude of association between mCAIDE and dementia, in comparison to CAIDE, mCAIDE was significantly associated with an increased risk of dementia in Black participants. These findings are consistent with previous studies comparing the predictive ability of different dementia risk scores. In cohorts from LMIC, dementia risk scores including the Australian National University Alzheimer’s Disease Risk Index (ANU-ADRI; c = 0.66–0.78); the Brief Dementia Screening Indicator (BDSI; c = 0.62–0.78); and the Rotterdam Study Basic Dementia Risk Model (BDRM; c = 0.66–0.78) showed similar levels of discriminative ability to that of the original development cohort, and where higher than that of CAIDE [10]. Furthermore, the magnitude of the association of CAIDE with global cognition was smaller than that of the Washington Heights-Inwood Columbia Aging Project (WHICAP) dementia risk score, which includes ethnicity in its calculation [31]. The strength of WHICAP with global cognition also did not differ between groups.

As the strongest genetic risk factor for late-onset AD, *APOE* displays ancestry-specific effects that may be due to gene-environment interactions [16,17]. In the Cardiovascular Risk Factors, Aging, and Incidence of Dementia (CAIDE) study of middle-aged Finnish individuals (n = 1,449), unfavorable risk profiles (physical activity, diet, smoking, alcohol intake) were associated with increased risk of incident dementia in *APOE*\*ε4 carriers only [18]. In contrast, in older adults from the Rotterdam study (n = 6,352), unfavorable risk profiles (smoking, depression, diabetes, physical activity, social isolation, and diet) were associated with increased incident dementia in *APOE*\*e4 non-carriers only [23]. In the multi-ethnic Washington Heights-Inwood Columbia Aging Project (WHICAP, n = 1,987, 28% NLW, 29% Black, 40% Latinx), using Life’s Simple 7 (LS7) – a risk score composed of physical activity, smoking, BMI, diet, cholesterol, blood glucose, and blood pressure used to improve cardiovascular health and reduce the risk of heart disease – better cardiovascular health was associated with reduced incidence of dementia in elderly *APOE*\*ε4 non-carriers only [22]. However, in the Atherosclerosis Risk in Communities Study (ARIC, n = 13,715, 75% NLW, 25% Black), better cardiovascular health as measured using LS7 was associated with lower incidence rates of dementia in ε4 non-carriers compared to ε4 carriers [21]. In sex- and race-stratified analyses, a significant interaction was observed in women such that there was a stronger association between cardiovascular health and dementia in *APOE*\*ε4 non-carriers. No interactions were observed in the whole cohort or other subpopulations. Finally, in the Chicago Health and Aging Project (CHAP, n = 3,886, 60% Black, 40% White), adherence to a healthy lifestyle (diet, cognitively stimulating activities, non-smoker, physical activity, light-moderate alcohol intake), was associated with slower cognitive decline in both *APOE*\*ε4 carriers and non-carriers [19]. In race-stratified analyses, the protective effect of a healthy lifestyle was stronger in NLW participants than in Blacks [20].

Together, these previous studies, in addition to our results, highlight that unfavorable risk factor profiles moderate the effect of *APOE* on dementia and cognitive impairment. However, the sample and methodological characteristics of each study introduce uncertainty on whether these effects are observed in *APOE*\*ε4 carriers, non-carriers, or both. In particular, the composition and weighting of the risk scores used, whether the risk factors are measured in mid-life or older age, sex- and race/ethnic-specific effects, and neuropathological heterogeneity in clinical AD diagnosis may affect the observed associations. As such, if dementia risk scores are to be used in precision medicine approaches for risk prediction and stratification, it is crucial to evaluate their generalizability across diverse populations.

Our study has several limitations. First, our findings are limited by the disproportionate sample sizes: NLW participants outnumber Blacks tenfold and Latinx and Asians twentyfold, impacting statistical power and the feasibility of longitudinal modeling. Second, the cross-sectional design precludes examining the association of CAIDE/mCAIDE risk scores with incident dementia. In particular, since CAIDE was designed to predict the midlife risk of dementia among individuals aged 45-60, and the mean age of NACC/ADNI participants is 72, the contribution of age to CAIDE in the NACC/ADNI cohort is underestimated, as the majority of participants are aged 65 or older. Third, the use of broad US Census racial/ethnic categories may overlook within-group heterogeneity, especially among Asian and Latinx populations. Fourth, the clinical nature of NACC and ADNI may affect the generalizability of our results to the general population. Fifth, the lack of comprehensive data on lifestyle factors and social determinants of health in these datasets precludes using more comprehensive dementia risk scores. Due to this, we were also unable to include physical activity in the CAIDE/mCAIDE risk scores, however, CAIDE remains predictive of dementia when physical activity is not included [26]. Finally, while *APOE* genotype is the strongest genetic risk factor for late-onset AD, a further 80+ loci are associated with AD [33]. As such, further work is needed to evaluate how lifestyle risk factors moderate the genetic liability for AD using cross-ancestry polygenic risk scores. Despite these limitations, our work addresses a significant gap in the literature by evaluating the influence of race/ethnicity on the effect of dementia risk scores and *APOE* on dementia risk.

In summary, using a large multi-ethnic cohort, we found that the CAIDE risk score, which was developed in a homogeneous population, exhibited race/ethnic-specific associations with dementia and notably was not associated with dementia risk among Black Americans. In contrast, a modified CAIDE risk score that was recalibrated based on a multi-ethnic cohort, was associated with increased dementia risk in Asian, Black, Latinx, and NLW Americans. Furthermore, unfavorable risk profiles were observed to exacerbate the risk effect of *APOE*\*e4 and attenuate the protective effect of *APOE*\*e2 in NLW and Blacks. These findings underscore the necessity of evaluating the validity of dementia risk scores in diverse populations for their effective integration into precision medicine strategies to promote brain health.

## Data Availability

All data produced in the present study are available upon reasonable request to the authors.

https://naccdata.org/

https://adni.loni.usc.edu/

## NACC

The NACC database is funded by NIA/NIH Grant U24 AG072122. NACC data are contributed by the NIA-funded ADRCs: P30 AG062429 (PI James Brewer, MD, PhD), P30 AG066468 (PI Oscar Lopez, MD), P30 AG062421 (PI Bradley Hyman, MD, PhD), P30 AG066509 (PI Thomas Grabowski, MD), P30 AG066514 (PI Mary Sano, PhD), P30 AG066530 (PI Helena Chui, MD), P30 AG066507 (PI Marilyn Albert, PhD), P30 AG066444 (PI John Morris, MD), P30 AG066518 (PI Jeffrey Kaye, MD), P30 AG066512 (PI Thomas Wisniewski, MD), P30 AG066462 (PI Scott Small, MD), P30 AG072979 (PI David Wolk, MD), P30 AG072972 (PI Charles DeCarli, MD), P30 AG072976 (PI Andrew Saykin, PsyD), P30 AG072975 (PI David Bennett, MD), P30 AG072978 (PI Neil Kowall, MD), P30 AG072977 (PI Robert Vassar, PhD), P30 AG066519 (PI Frank LaFerla, PhD), P30 AG062677 (PI Ronald Petersen, MD, PhD), P30 AG079280 (PI Eric Reiman, MD), P30 AG062422 (PI Gil Rabinovici, MD), P30 AG066511 (PI Allan Levey, MD, PhD), P30 AG072946 (PI Linda Van Eldik, PhD), P30 AG062715 (PI Sanjay Asthana, MD, FRCP), P30 AG072973 (PI Russell Swerdlow, MD), P30 AG066506 (PI Todd Golde, MD, PhD), P30 AG066508 (PI Stephen Strittmatter, MD, PhD), P30 AG066515 (PI Victor Henderson, MD, MS), P30 AG072947 (PI Suzanne Craft, PhD), P30 AG072931 (PI Henry Paulson, MD, PhD), P30 AG066546 (PI Sudha Seshadri, MD), P20 AG068024 (PI Erik Roberson, MD, PhD), P20 AG068053 (PI Justin Miller, PhD), P20 AG068077 (PI Gary Rosenberg, MD), P20 AG068082 (PI Angela Jefferson, PhD), P30 AG072958 (PI Heather Whitson, MD), P30 AG072959 (PI James Leverenz, MD).

## ADNI

**\*** Data used in preparation of this article were obtained from the Alzheimer’s Disease Neuroimaging Initiative (ADNI) database (adni.loni.usc.edu). As such, the investigators within the ADNI contributed to the design and implementation of ADNI and/or provided data but did not participate in analysis or writing of this report. A complete listing of ADNI investigators can be found at: http://adni.loni.usc.edu/wp-content/uploads/how_to_apply/ADNI_Acknowledgement_List.pdf

Data collection and sharing for this project was funded by the Alzheimer’s Disease Neuroimaging Initiative (ADNI) (National Institutes of Health Grant U01 AG024904) and DOD ADNI (Department of Defense award number W81XWH-12-2-0012). ADNI is funded by the National Institute on Aging, the National Institute of Biomedical Imaging and Bioengineering, and through generous contributions from the following: AbbVie, Alzheimer’s Association; Alzheimer’s Drug Discovery Foundation; Araclon Biotech; BioClinica, Inc.; Biogen; Bristol-Myers Squibb Company; CereSpir, Inc.; Cogstate; Eisai Inc.; Elan Pharmaceuticals, Inc.; Eli Lilly and Company; EuroImmun; F. Hoffmann-La Roche Ltd and its affiliated company Genentech, Inc.; Fujirebio; GE Healthcare; IXICO Ltd.;Janssen Alzheimer Immunotherapy Research & Development, LLC.; Johnson & Johnson Pharmaceutical Research & Development LLC.; Lumosity; Lundbeck; Merck & Co., Inc.;Meso Scale Diagnostics, LLC.; NeuroRx Research; Neurotrack Technologies; Novartis Pharmaceuticals Corporation; Pfizer Inc.; Piramal Imaging; Servier; Takeda Pharmaceutical Company; and Transition Therapeutics. The Canadian Institutes of Health Research is providing funds to support ADNI clinical sites in Canada. Private sector contributions are facilitated by the Foundation for the National Institutes of Health (www.fnih.org). The grantee organization is the Northern California Institute for Research and Education, and the study is coordinated by the Alzheimer’s Therapeutic Research Institute at the University of Southern California. ADNI data are disseminated by the Laboratory for Neuro Imaging at the University of Southern California.

## Author Contributions

Dr. Andrews had full access to all the data in the study and takes full responsibility for the integrity of the data and the accuracy of the data analysis. The code to support the analysis of this study is available at: https://github.com/AndrewsLabUCSF/CAIDE_APOE.git

Concept and design: SJA, MEB, AER, BFH, WDB, KY

Acquisition, analysis, or interpretation of data: SJA, AIB

Drafting of the Manuscript: SJA, KY, AIB

Critical review of the manuscript for important intellectual content: MEB, AER, BFH, WDB, KY

Statistical analysis: SJA, AIB

## Consent Statement

Participants provided informed consent, and institutional review board approval was locally obtained.

## References

[1] 2023 Alzheimer’s disease facts and figures. Alzheimer’s Dement 2023;19:1598–695.

[2] Frisoni GB, Altomare D, Ribaldi F, Villain N, Brayne C, Mukadam N, et al. Dementia prevention in memory clinics: recommendations from the European task force for brain health services. Lancet Regional Heal - Europe 2023:100576.

[3] Widén E, Junna N, Ruotsalainen S, Surakka I, Mars N, Ripatti P, et al. How Communicating Polygenic and Clinical Risk for Atherosclerotic Cardiovascular Disease Impacts Health Behavior: an Observational Follow-up Study. Circulation Genom Precis Medicine 2022;15:e003459.

[4] Anstey KJ, Zheng L, Peters R, Kootar S, Barbera M, Stephen R, et al. Dementia Risk Scores and Their Role in the Implementation of Risk Reduction Guidelines. Front Neurol 2022;12:765454.

[5] Mindt MR, Okonkwo O, Weiner MW, Veitch DP, Aisen P, Ashford M, et al. Improving generalizability and study design of Alzheimer’s disease cohort studies in the United States by including under-represented populations. Alzheimer’s Dementia 2022.

[6] Barnes DE, Yaffe K. The projected effect of risk factor reduction on Alzheimer’s disease prevalence. Lancet Neurol 2011;10:819--828.

[7] Livingston G, Huntley J, Liu KY, Costafreda SG, Selbæk G, Alladi S, et al. Dementia prevention, intervention, and care: 2024 report of the Lancet standing Commission. Lancet 2024.

[8] Kivipelto M, Ngandu T, Laatikainen T, Winblad B, Soininen H, Tuomilehto J. Risk score for the prediction of dementia risk in 20 years among middle aged people: a longitudinal, population-based study. Lancet Neurology 2006;5:735–41.

[9] Ngandu T, Lehtisalo J, Solomon A, Levälahti E, Ahtiluoto S, Antikainen R, et al. A 2 year multidomain intervention of diet, exercise, cognitive training, and vascular risk monitoring versus control to prevent cognitive decline in at-risk elderly people (FINGER): a randomised controlled trial. Lancet 2015;385:2255–63.

[10] Stephan BCM, Pakpahan E, Siervo M, Licher S, Muniz-Terrera G, Mohan D, et al. Prediction of dementia risk in low-income and middle-income countries (the 10/66 Study): an independent external validation of existing models. Lancet Global Heal 2020;8:e524–35.

[11] Schaich CL, Yeboah J, Espeland MA, Baker LD, Ding J, Hayden KM, et al. Association of Vascular Risk Scores and Cognitive Performance in a Diverse Cohort: The Multi-Ethnic Study of Atherosclerosis. Journals Gerontology Ser 2021;77:1208–15.

[12] Tolea MI, Heo J, Chrisphonte S, Galvin JE. A Modified CAIDE Risk Score as a Screening Tool for Cognitive Impairment in Older Adults. J Alzheimer’s Dis 2021;82:1755–68.

[13] Reiman EM, Arboleda-Velasquez JF, Quiroz YT, Huentelman MJ, Beach TG, Caselli RJ, et al. Exceptionally low likelihood of Alzheimer’s dementia in APOE2 homozygotes from a 5,000-person neuropathological study. Nat Commun 2020;11:667.

[14] Farrer LA, Cupples LA, Haines JL, Hyman B, Kukull WA, Mayeux R, et al. Effects of Age, Sex, and Ethnicity on the Association Between Apolipoprotein E Genotype and Alzheimer Disease: A Meta-analysis. Jama 1997;278:1349–56.

[15] Belloy ME, Andrews SJ, Guen YL, Cuccaro M, Farrer LA, Napolioni V, et al. APOE Genotype and Alzheimer Disease Risk Across Age, Sex, and Population Ancestry. JAMA Neurol 2023;80:1284–94.

[16] Raine A. Biosocial Studies of Antisocial and Violent Behavior in Children and Adults: A Review. J Abnorm Child Psychol 2002;30:311–26.

[17] Boardman JD, Domingue BW, Blalock CL, Haberstick BC, Harris KM, McQueen MB. Is the Gene-Environment Interaction Paradigm Relevant to Genome-Wide Studies? The Case of Education and Body Mass Index. Demography 2014;51:119–39.

[18] Kivipelto M, Rovio S, Ngandu T, Kåreholt I, Eskelinen M, Winblad B, et al. Apolipoprotein E lll4 magnifies lifestyle risks for dementia: a population-based study. J Cell Mol Med 2008;12:2762–71.

[19] Dhana K, Aggarwal NT, Rajan KB, Barnes LL, Evans DA, Morris MC. Impact of the Apolipoprotein E4 allele on the Relationship Between Healthy Lifestyle and Cognitive Decline: A Population-based Study. Am J Epidemiology 2021;190:kwab033.

[20] Dhana K, Barnes LL, Liu X, Agarwal P, Desai P, Krueger KR, et al. Genetic risk, adherence to a healthy lifestyle, and cognitive decline in African Americans and European Americans. Alzheimer’s Dement 2022;18:572–80.

[21] Lee M, Hughes TM, George KM, Griswold ME, Sedaghat S, Simino J, et al. Education and Cardiovascular Health as Effect Modifiers of APOE ε4 on Dementia: The Atherosclerosis Risk in Communities Study. J Gerontol: Ser A 2021;77:1199–207.

[22] Guo J, Brickman AM, Manly JJ, Reitz C, Schupf N, Mayeux RP, et al. Association of Life’s Simple 7 with incident dementia and its modification by the apolipoprotein E genotype. Alzheimer’s Dementia 2021;17:1905–13.

[23] Licher S, Ahmad S, Karamujić-Čomić H, Voortman T, Leening MJG, Ikram MA, et al. Genetic predisposition, modifiable-risk-factor profile and long-term dementia risk in the general population. Nat Med 2019;25:1364–9.

[24] Beekly DL, Ramos EM, Lee WW, Deitrich WD, Jacka ME, Wu J, et al. The National Alzheimer’s Coordinating Center (NACC) Database: The Uniform Data Set. Alzheimer Dis Assoc Disord 2007;21:249–58.

[25] Petersen RC, Aisen PS, Beckett LA, Donohue MC, Gamst AC, Harvey DJ, et al. Alzheimer’s Disease Neuroimaging Initiative (ADNI) Clinical characterization. Neurology 2010;74:201–9.

[26] Exalto LG, Quesenberry CP, Barnes D, Kivipelto M, Biessels GJ, Whitmer RA. Midlife risk score for the prediction of dementia four decades later. Alzheimer’s Dementia J Alzheimer’s Assoc 2013;10:562–70.

[27] Stekhoven DJ, Bühlmann P. MissForest—non-parametric missing value imputation for mixed-type data. Bioinformatics 2012;28:112–8.

[28] Saykin AJ, Shen L, Foroud TM, Potkin SG, Swaminathan S, Kim S, et al. Alzheimer’s Disease Neuroimaging Initiative biomarkers as quantitative phenotypes: Genetics core aims, progress, and plans. Alzheimer’s Dement 2010;6:265–73.

[29] Naj AC, Jun G, Beecham GW, Wang L-S, Vardarajan BN, Buros J, et al. Common variants at MS4A4/MS4A6E, CD2AP, CD33 and EPHA1 are associated with late-onset Alzheimer’s disease. Nat Genet 2011;43:436–41.

[30] Paternoster R, Brame R, Mazerolle P, Piquero A. USING THE CORRECT STATISTICAL TEST FOR THE EQUALITY OF REGRESSION COEFFICIENTS. Criminology 1998;36:859–66.

[31] Torres S, Alexander A, O’Bryant S, Medina LD. Cognition and the Predictive Utility of Three Risk Scores in an Ethnically Diverse Sample. J Alzheimer’s Dis 2020;75:1049–59.

[32] Nianogo RA, Rosenwohl-Mack A, Yaffe K, Carrasco A, Hoffmann CM, Barnes DE. Risk Factors Associated With Alzheimer Disease and Related Dementias by Sex and Race and Ethnicity in the US. Jama Neurol 2022;79:584–91.

[33] Andrews SJ, Renton AE, Fulton-Howard B, Podlesny-Drabiniok A, Marcora E, Goate AM. The complex genetic architecture of Alzheimer’s disease: novel insights and future directions. EBioMedicine 2023;90:104511.

