## Supplementary Figure1; Tables 1-2 for "Dementia Risk Scores, *APOE,* and risk of Alzheimer disease: one size does not fit all"

**Supplements**

**Supplementary Figures**

**Supplementary Figure 1:**

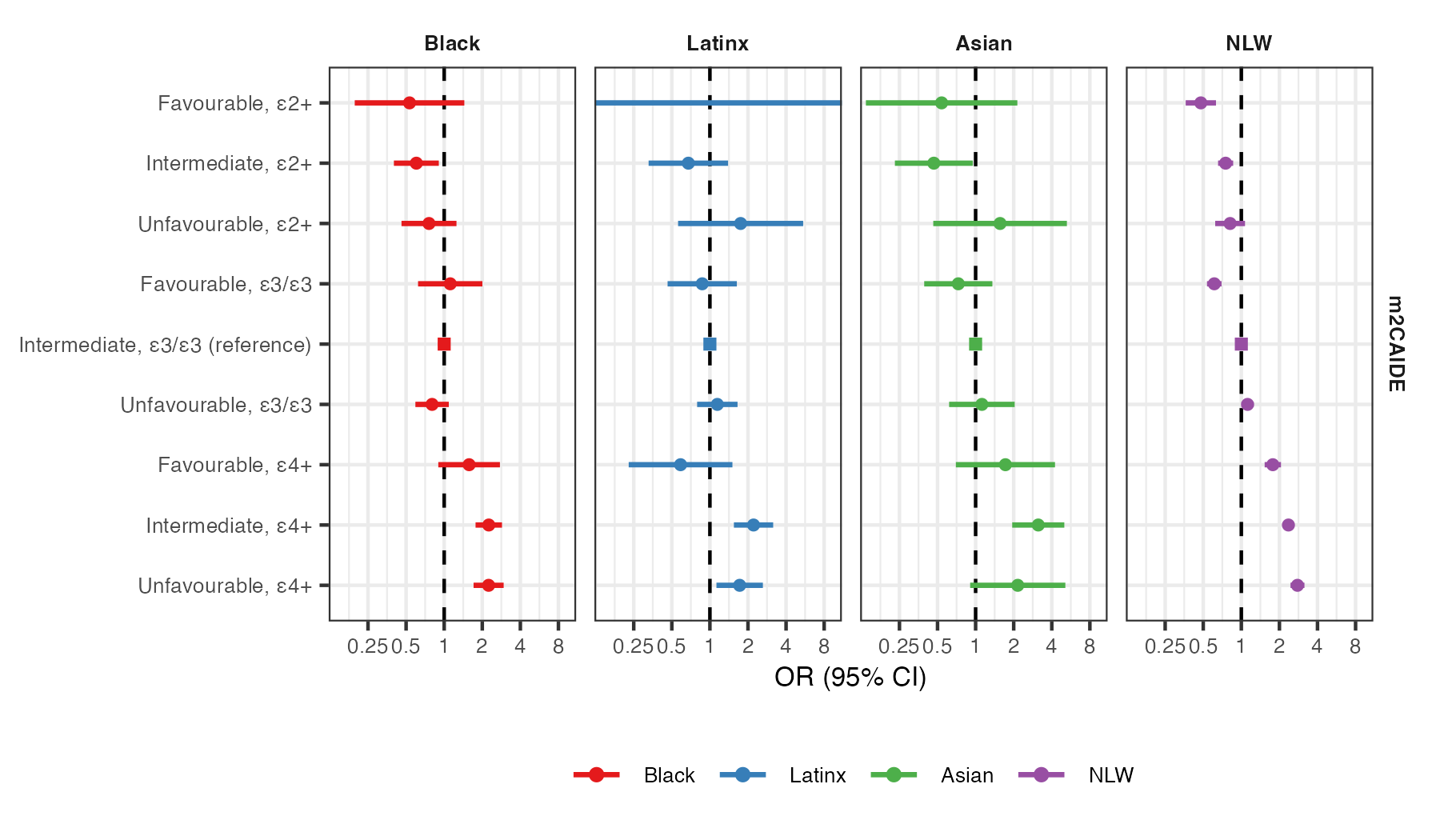

**Supplementary Table 1: Risk of ADRD/MCI according to genetic and modifiable risk factor burden.** A modified mCAIDE (m^2^CAIDE) score was constructed composed only of modifiable risk factors - education, hypertension, obesity, and hypercholesteremia. m^2^CAIDE was categorized into tertiles representing favorable (m^2^CAIDE < 1), intermediate (m^2^CAIDE >= 1 & < 5), and unfavorable (m^2^CAIDE >=5 & < 9) risk profiles. Intermediate risk profiles and *APOE* ε3/ε3 were used as the reference category.

**Supplementary Tables**

**Supplementary Table 1:** CAIDE scoring algorithm and cohort description

**Supplementary Table 2:** mCAIDE scoring algorithm and cohort description

**Supplementary Table 3:** Summary of Baseline Characteristics of Joint NACC and ADNI

**Supplementary Table 4:** Person's Chi-squared test for racial/ethnic group differences in gender, hypercholesterolemia, hypertension, ADRD/MCI diagnosis, and APOE e4

**Supplementary Table 5:** Analysis of Variance (ANOVA) for racial/ethnic group differences in age, education, BMI, CAIDE, and mCAIDE.

**Supplementary Table 6:** The association of *APOE* and CAIDE with dementia and cognitive impairment stratified by race/ethnicity.

**Supplementary Table 7:** Comparison of the association of APOE genotype and CAIDE with dementia and cognitive impairment between racial/ethnic groups.

**Supplementary Table 8:** Delong's Test Results for Comparing the Discriminative Power of Predictive Models stratified by race/ethnicity

**Supplementary Table 9:** Association of CAIDE and APOE on MCI/ADRD stratified by Sex

**Supplementary Table 10:** Association of CAIDE and APOE with ADRD/MCI stratified by sex and race/ethnicity

**Supplementary Table 11:** Association of individual risk factors (age, education, gender, BMI, hypercholesteremia, systolic BP) on MCI and ADRD stratified by race/ethnicity

**Supplementary Table 12:** The association of APOE and mCAIDE with dementia and cognitive impairment stratified by race/ethnicity.

**Supplementary Table 13:** Comparison of the association of APOE genotype and mCAIDE with dementia and cognitive impairment between racial/ethnic groups.

**Supplementary Table 14:** Association of mCAIDE and APOE on MCI/ADRD stratified by Sex.

**Supplementary Table 15:** Association of mCAIDE and APOE with ADRD/MCI stratified by sex and race/ethnicity.

**Supplementary Table 16:** The association of CAIDE and APOE interaction on MCI/ADRD stratified by race/ethnicity

**Supplementary Table 17:** The association of mCAIDE and APOE interaction on MCI/ADRD stratified by race/ethnicity

**Supplementary Table 18:** Association of m^2^CAIDE and APOE on MCI/ADRD stratified by race/ethnicity

**Supplementary Table 19:** Comparison of the association of combined APOE genotype and CAIDE risk profiles with dementia and cognitive impairment between racial/ethnic groups.

**Supplementary Table 20:** Comparison of the association of combined APOE genotype and mCAIDE risk profiles with dementia and cognitive impairment between racial/ethnic groups.

**Table S1: CAIDE scoring algorithm and cohort description**

|  | **CAIDE**  **Score** | **Asian**  N = 573*^1^* | **Black**  N = 2,259*^1^* | **Latinx**  N = 961*^1^* | **NLW**  N = 16,962*^1^* |
| --- | --- | --- | --- | --- | --- |
| **Age** |  |  |  |  |  |
| <47 | 0 | **-** | **-** | **-** | **-** |
| 47-53 | 3 | **-** | **-** | **-** | **-** |
| >53 | 4 | 573 (100%) | 2,259 (100%) | 961 (100%) | 16,962 (100%) |
| **Gender** |  |  |  |  |  |
| Female | 0 | 324 (57%) | 1,647 (73%) | 614 (64%) | 9,076 (54%) |
| Male | 1 | 249 (43%) | 612 (27%) | 347 (36%) | 7,886 (46%) |
| **Education** |  |  |  |  |  |
| ≥10 | 0 | 544 (95%) | 2,127 (94%) | 739 (77%) | 16,751 (99%) |
| 7-9 | 2 | 12 (2.1%) | 90 (4.0%) | 78 (8.1%) | 176 (1.0%) |
| <7 | 3 | 17 (3.0%) | 42 (1.9%) | 144 (15%) | 35 (0.2%) |
| **BMI** |  |  |  |  |  |
| <30 | 0 | 536 (94%) | 1,343 (59%) | 709 (74%) | 13,645 (80%) |
| ≥30 | 2 | 37 (6.5%) | 916 (41%) | 252 (26%) | 3,317 (20%) |
| **Hypertension** |  |  |  |  |  |
| No | 0 | 369 (64%) | 1,250 (55%) | 566 (59%) | 10,947 (65%) |
| Yes | 2 | 204 (36%) | 1,009 (45%) | 395 (41%) | 6,015 (35%) |
| **Hypercholesterolemia** |  |  |  |  |  |
| No | 0 | 292 (51%) | 1,145 (51%) | 462 (48%) | 9,310 (55%) |
| Yes | 2 | 281 (49%) | 1,114 (49%) | 499 (52%) | 7,652 (45%) |
| **CAIDE** |  | 6.39 (1.71) | 7.10 (1.90) | 7.36 (2.31) | 6.49 (1.79) |
| **CAIDE Missing** |  |  |  |  |  |
| 0 |  | 445 (78%) | 1,836 (81%) | 784 (82%) | 13,936 (82%) |
| 1 |  | 93 (16%) | 347 (15%) | 135 (14%) | 2,244 (13%) |
| 2 |  | 30 (5.2%) | 73 (3.2%) | 39 (4.1%) | 732 (4.3%) |
| 3 |  | 5 (0.9%) | 3 (0.1%) | 3 (0.3%) | 50 (0.3%) |
| **CAIDE** |  |  |  |  |  |
| High |  | 69 (12%) | 462 (20%) | 282 (29%) | 2,644 (16%) |
| Mid |  | 399 (70%) | 1,526 (68%) | 553 (58%) | 11,392 (67%) |
| Low |  | 105 (18%) | 271 (12%) | 126 (13%) | 2,926 (17%) |
| **CAIDE x *APOE*** |  |  |  |  |  |
| High, ε2+ |  | 8 (1.4%) | 52 (2.3%) | 10 (1.0%) | 181 (1.1%) |
| High, ε3/e3 |  | 44 (7.7%) | 191 (8.5%) | 164 (17%) | 1,369 (8.1%) |
| High, ε4+ |  | 17 (3.0%) | 219 (9.7%) | 108 (11%) | 1,094 (6.4%) |
| Mid, ε2+ |  | 45 (7.9%) | 190 (8.4%) | 26 (2.7%) | 1,024 (6.0%) |
| Mid, ε3/e3 |  | 237 (41%) | 629 (28%) | 325 (34%) | 5,519 (33%) |
| Mid, ε4+ |  | 117 (20%) | 707 (31%) | 202 (21%) | 4,849 (29%) |
| Low, ε2+ |  | 13 (2.3%) | 42 (1.9%) | 13 (1.4%) | 321 (1.9%) |
| Low, ε3/e3 |  | 64 (11%) | 96 (4.2%) | 73 (7.6%) | 1,497 (8.8%) |
| Low, ε 4+ |  | 28 (4.9%) | 133 (5.9%) | 40 (4.2%) | 1,108 (6.5%) |

**Table S2: mCAIDE scoring algorithm and cohort description**

|  | **mCAIDE**  **Score** | **Asian**  N = 573*^1^* | **Black**  N = 2,259*^1^* | **Latinx**  N = 961*^1^* | **NLW**  N = 16,962*^1^* |
| --- | --- | --- | --- | --- | --- |
| **Age** |  |  |  |  |  |
| <65 | 0 | 122 (21%) | 367 (16%) | 159 (17%) | 2,699 (16%) |
| 56-72 | 1 | 173 (30%) | 825 (37%) | 347 (36%) | 5,528 (33%) |
| >73 | 2 | 278 (49%) | 1,067 (47%) | 455 (47%) | 8,735 (51%) |
| **Gender** |  |  |  |  |  |
| Female | 0 | 324 (57%) | 1,647 (73%) | 614 (64%) | 9,076 (54%) |
| Male | 1 | 249 (43%) | 612 (27%) | 347 (36%) | 7,886 (46%) |
| **Education** |  |  |  |  |  |
| ≥16 | 0 | 237 (41%) | 577 (26%) | 207 (22%) | 6,754 (40%) |
| 12-16 | 1 | 296 (52%) | 1,429 (63%) | 492 (51%) | 9,799 (58%) |
| <12 | 2 | 40 (7.0%) | 253 (11%) | 262 (27%) | 409 (2.4%) |
| **BMI** |  |  |  |  |  |
| <30 | 0 | 536 (94%) | 1,343 (59%) | 709 (74%) | 13,645 (80%) |
| ≥30 | 2 | 37 (6.5%) | 916 (41%) | 252 (26%) | 3,317 (20%) |
| **Hypertension** |  |  |  |  |  |
| No | 0 | 369 (64%) | 1,250 (55%) | 566 (59%) | 10,947 (65%) |
| Yes | 2 | 204 (36%) | 1,009 (45%) | 395 (41%) | 6,015 (35%) |
| **Hypercholesterolemia** |  |  |  |  |  |
| No | 0 | 292 (51%) | 1,145 (51%) | 462 (48%) | 9,310 (55%) |
| Yes | 2 | 281 (49%) | 1,114 (49%) | 499 (52%) | 7,652 (45%) |
| **mCAIDE** |  | 4.18 (2.01) | 5.12 (2.13) | 5.10 (2.28) | 4.44 (2.06) |
| **mCAIDE Missing** |  |  |  |  |  |
| 0 |  | 445 (78%) | 1,836 (81%) | 784 (82%) | 13,936 (82%) |
| 1 |  | 93 (16%) | 347 (15%) | 135 (14%) | 2,244 (13%) |
| 2 |  | 30 (5.2%) | 73 (3.2%) | 39 (4.1%) | 732 (4.3%) |
| 3 |  | 5 (0.9%) | 3 (0.1%) | 3 (0.3%) | 50 (0.3%) |
| **mCAIDE** |  |  |  |  |  |
| High |  | 77 (13%) | 609 (27%) | 272 (28%) | 2,875 (17%) |
| Mid |  | 369 (64%) | 1,393 (62%) | 566 (59%) | 10,987 (65%) |
| Low |  | 127 (22%) | 257 (11%) | 123 (13%) | 3,100 (18%) |
| **mCAIDE x *APOE*** |  |  |  |  |  |
| High, ε2+ |  | 10 (1.7%) | 68 (3.0%) | 11 (1.1%) | 198 (1.2%) |
| High, ε3/e3 |  | 49 (8.6%) | 249 (11%) | 162 (17%) | 1,498 (8.8%) |
| High, ε4+ |  | 18 (3.1%) | 292 (13%) | 99 (10%) | 1,179 (7.0%) |
| Mid, ε2+ |  | 41 (7.2%) | 175 (7.7%) | 28 (2.9%) | 995 (5.9%) |
| Mid, ε3/e3 |  | 217 (38%) | 572 (25%) | 328 (34%) | 5,325 (31%) |
| Mid, ε4+ |  | 111 (19%) | 646 (29%) | 210 (22%) | 4,667 (28%) |
| Low, ε2+ |  | 15 (2.6%) | 41 (1.8%) | 10 (1.0%) | 333 (2.0%) |
| Low, ε3/e3 |  | 79 (14%) | 95 (4.2%) | 72 (7.5%) | 1,562 (9.2%) |
| Low, ε 4+ |  | 33 (5.8%) | 121 (5.4%) | 41 (4.3%) | 1,205 (7.1%) |
